# A Retrospective Longitudinal Study of COVID-19 as Seen by a Large Urban Hospital in Chicago

**DOI:** 10.1101/2020.11.29.20240606

**Authors:** Haotian Chen, Yogatheesan Varatharajah, Sarah Stewart de Ramirez, Paul Arnold, Casey Frankenberger, Bala Hota, Ravishankar Iyer

## Abstract

The rapid spread of the novel coronavirus disease 2019 (COVID-19) has created high demand for medical resources, including personnel, intensive care unit beds, and ventilators. As thousands of patients are hospitalized, the disease has shown remarkable diversity in its manifestation; many patients with mild to no symptoms recover from the disease requiring minimal care, but some patients with severe disease progression require mechanical ventilation support in intensive care units (ICU) with an increased risk of death. Studying the characteristics of patients in these various strata can help us understand the varied progression of this disease, enable earlier interventions for at-risk patients, and help manage medical resources more efficiently. This paper presents a retrospective analysis of 10,123 COVID-19 patients treated at the Rush University Medical Center in Chicago, including their demographics, symptoms, comorbidities, laboratory values, vital signs, and clinical history. Specifically, we present a staging scheme based on discrete clinical events (i.e., admission to the hospital, admission to the ICU, mechanical ventilation, and death), and investigate the temporal trend of clinical variables and the effect of comorbidities in each of those stages. We then developed a prognostic model to predict ventilation demands at an individual patient level by analyzing baseline clinical variables, which entails (1) a least absolute shrinkage and selection operator (LASSO) regression and a decision tree model to identify predictors for mechanical ventilation; and (2) a logistic regression model based on these risk factors to predict which patients will eventually need ventilatory support. Our results indicate that the prognostic model achieves an AUC of 0.823 (95% CI: 0.765–0.880) in identifying patients who will eventually require mechanical ventilation.

## Introduction

Since the emergence of COVID-19, first identified in Wuhan, China [1], a global pandemic has ensued [2], and widespread, sustained transmission has been observed worldwide. As of this writing, the virus has infected ∼63 million people and caused over 1.4 million deaths worldwide, including over 13.75 million cases and over 273,000 deaths in the United States.

Studies have been published of the clinical characteristics and treatment outcomes in Chinese cities such as Wuhan [3], Shanghai [4], and Chongqing [5], as well as New York City [6,7]. These studies suggest that acute respiratory distress syndrome (ARDS) is a major driver of high mortality rates among critically ill patients. Several potentially impactful interventions have been found to reduce the severity of the illness and improve outcomes among critically ill patients with COVID-19: 1) early prone positioning (before intubation), which improves oxygenation and reduces the need for mechanical ventilation [8–10]; 2) Remdesivir therapy, when given to those requiring supplemental oxygen, can reduce recovery time [11]; and 3) dexamethasone, a frequent adjunctive therapy for ARDS and sepsis, has been found to reduce mortality among those with COVID-19 and respiratory compromise [12]. Those findings suggest the importance of early identification of ARDS as part of an overall COVID-19 treatment plan. Therefore, there is a critical need for a tool that can identify the patients who can benefit from earlier interventions.

In this single-center study conducted at the highest-volume clinical center for COVID-19 in the state of Illinois, we characterized the evolution of infections, risk factors for infection, and predictors of severe illness (i.e., ARDS), using a multistage analytic approach. Using LASSO regression and decision tree algorithms, we identified the key risk factors that predicted the need for mechanical ventilation from past data. Finally, we developed an early warning system for identifying those at high risk for requiring mechanical ventilation using the identified risk factors. An important aspect of the machine learning-based model is its ability to individualize a prediction based on features that are readily available upon hospital admission.

Although recently published comprehensive studies from China address disease progression, our study is unique as it focuses on the United States. We note that work in other urban areas of the United States (i.e., New York and Los Angeles) has found strong differences in severe illness onset and outcomes across racial and ethnic groups [13–22]. In that context, our research focuses specifically on the population of Chicago, Illinois, which has a diverse and unique urban patient population.

## Methods

The overall flow of our analysis is illustrated in Figure 1. Our analysis included 10,123 COVID-19 patients for whom information on demographics and initial symptoms was available. Among these patients, 1,788 were admitted to the hospital; for them, additional longitudinal information, such as lab test results and vitals, was available. Among the hospitalized patients, 301 had most of their lab measures and vitals collected on the day of admission. We used data collected from those patients to develop and validate the prediction model in a retrospective fashion. Out of the 301 investigated patients, 96 required ventilation. The predictive was developed in two steps: (1) we used a LASSO regression and a decision tree model to identify top risk factors for mechanical ventilation; and (2) we developed a logistic regression model based on these risk factors to predict which patients would eventually need ventilatory support.

**Figure 1.**
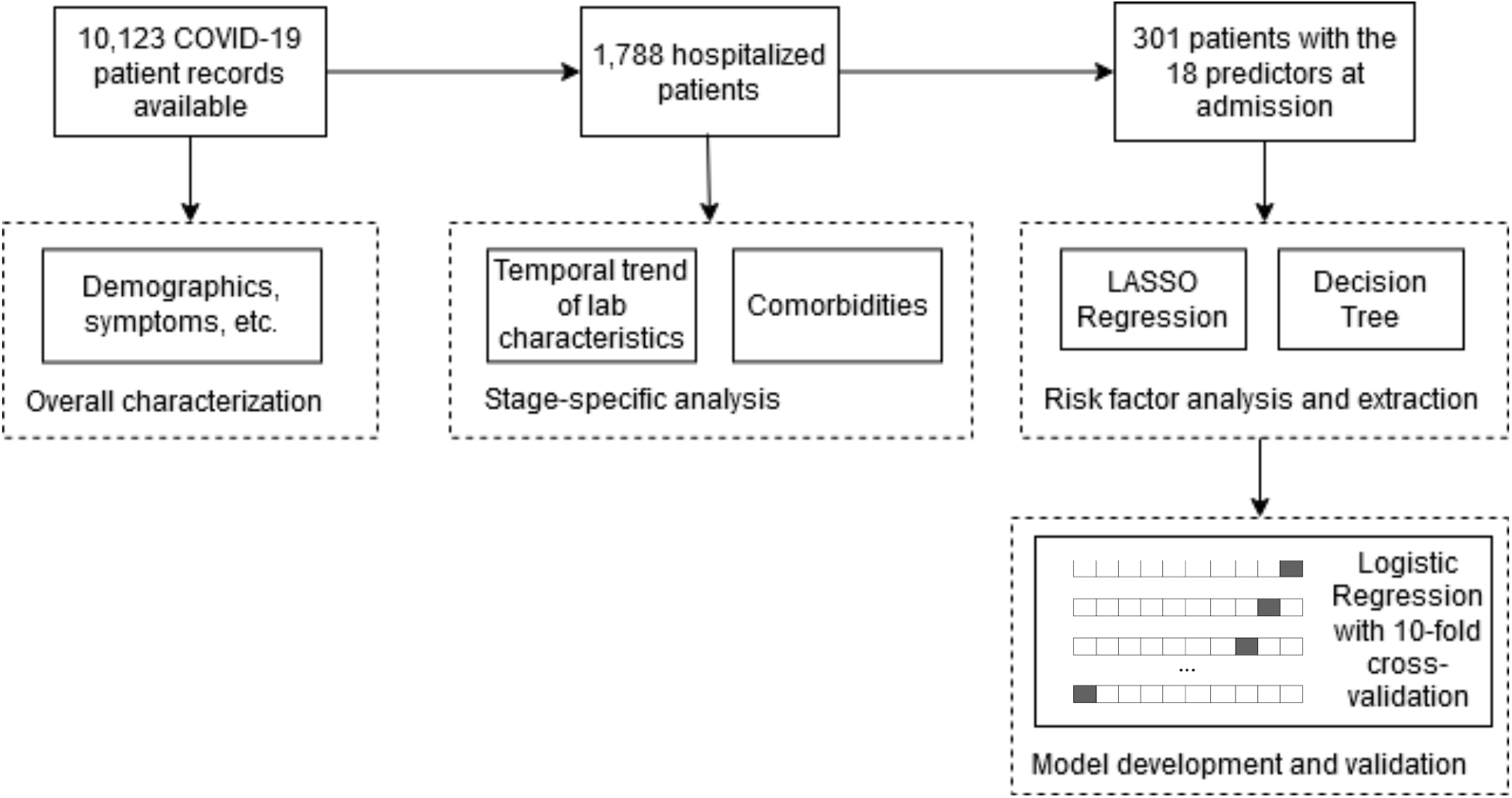
Overall flow of the analysis and the data: Our analysis included 10,123 COVID-19 patients, 1,788 of whom were admitted to the hospital. Among the hospitalized patients, 301 had most of their lab measures and vitals collected on the day of admission. We used their data to develop and validate the prediction model in a retrospective fashion. We developed the predictive model in two steps: (1) we used a LASSO regression and a decision tree model to identify key risk factors for mechanical ventilation; and (2) we developed a logistic regression model based on these risk factors to predict the need for ventilation support at a patient level.

### Data Sources

Data was collected at Rush University Medical Center in Chicago, Illinois, and cover the COVID-19 patients registered between 03/17/2020 and 08/07/2020. Information on patients and treatments was obtained from queries against data warehouses populated from regular exports of clinical data stored in Rush’s Epic electronic medical record (EMR) system. The study included 10,123 patients with COVID-19, and their EMRs were retrospectively processed to extract the patients’ demographics (Table 1), laboratory findings, vitals (Table 2), and comorbidities (Table 3). The age of each patient was typically documented as a numerical value, but for 41 patients whose ages were documented by the text “90+”, we assigned an age of 90 for use in our analysis.

**Table 1.**
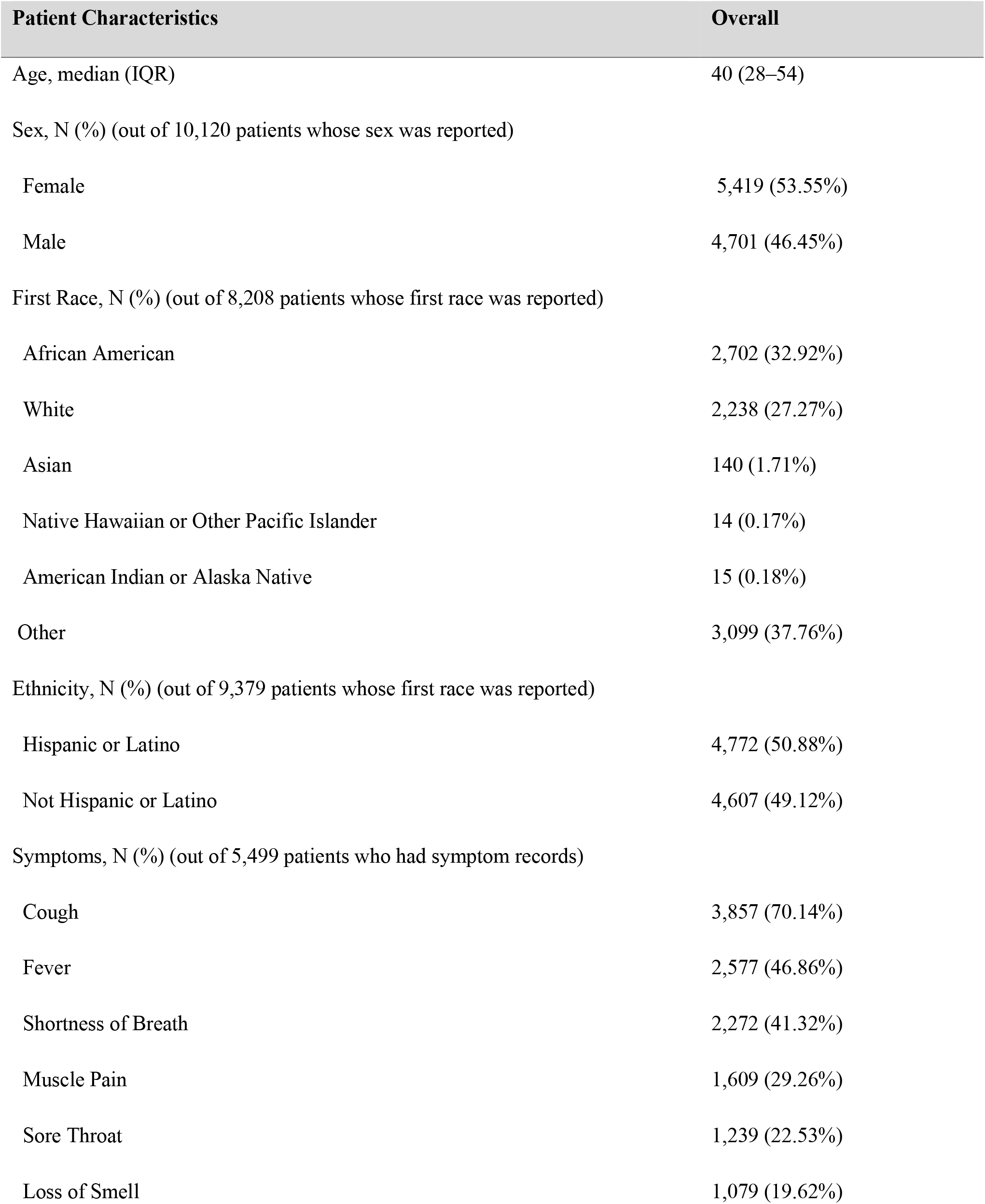
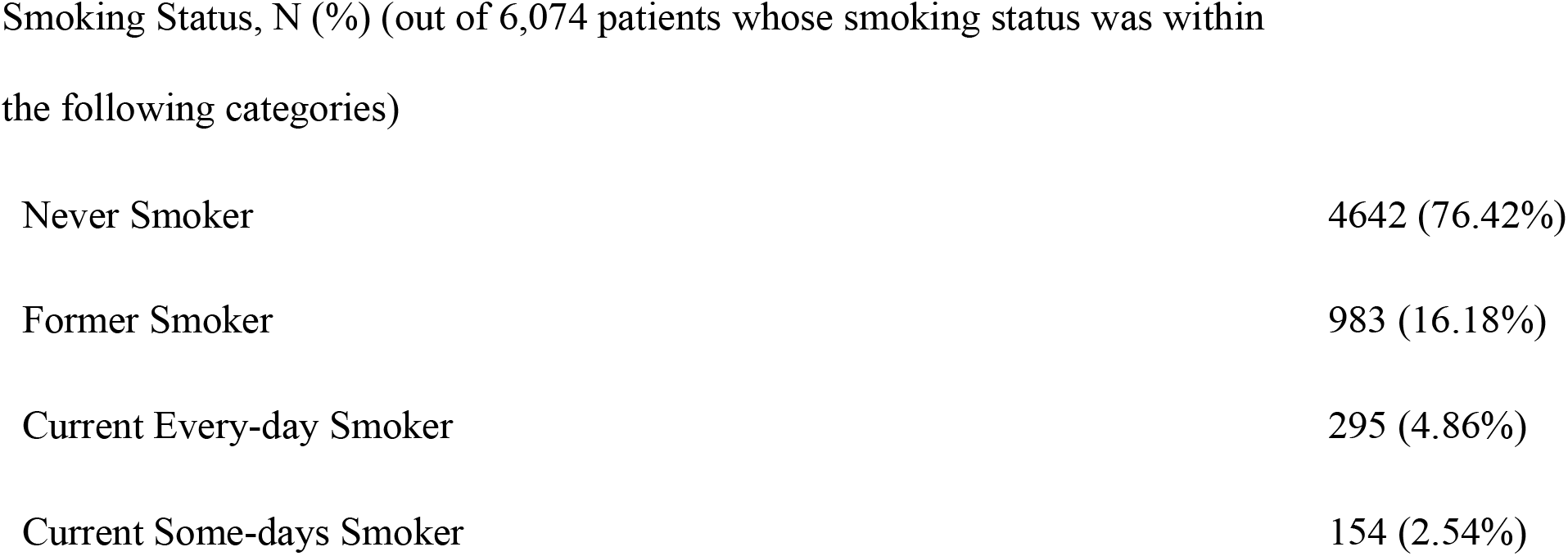
Demographics, symptoms, and smoking status of infected patients.

| Patient Characteristics | Overall |
| --- | --- |
| Age, median (IQR) | 40 (28–54) |
| Sex, N (%) (out of 10,120 patients whose sex was reported) |  |
| Female | 5,419 (53.55%) |
| Male | 4,701 (46.45%) |
| First Race, N (%) (out of 8,208 patients whose first race was reported) |  |
| African American | 2,702 (32.92%) |
| White | 2,238 (27.27%) |
| Asian | 140 (1.71%) |
| Native Hawaiian or Other Pacific Islander | 14 (0.17%) |
| American Indian or Alaska Native | 15 (0.18%) |
| Other | 3,099 (37.76%) |
| Ethnicity, N (%) (out of 9,379 patients whose first race was reported) |  |
| Hispanic or Latino | 4,772 (50.88%) |
| Not Hispanic or Latino | 4,607 (49.12%) |
| Symptoms, N (%) (out of 5,499 patients who had symptom records) |  |
| Cough | 3,857 (70.14%) |
| Fever | 2,577 (46.86%) |
| Shortness of Breath | 2,272 (41.32%) |
| Muscle Pain | 1,609 (29.26%) |
| Sore Throat | 1,239 (22.53%) |
| Loss of Smell | 1,079 (19.62%) |
| Smoking Status, N (%) (out of 6,074 patients whose smoking status was within the following categories) |  |
| Never Smoker | 4642 (76.42%) |
| Former Smoker | 983 (16.18%) |
| Current Every-day Smoker | 295 (4.86%) |
| Current Some-days Smoker | 154 (2.54%) |

**Table 2.**
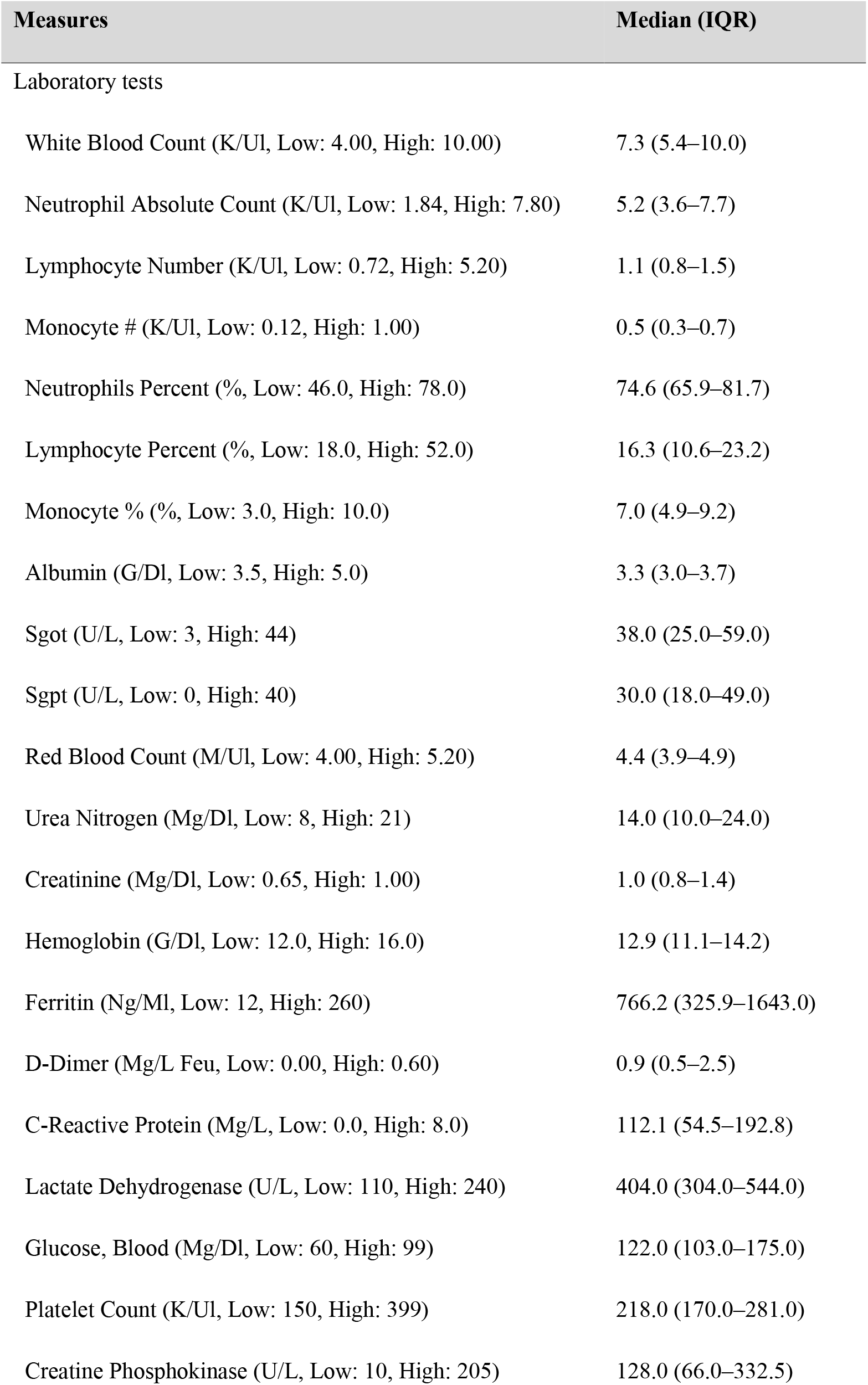
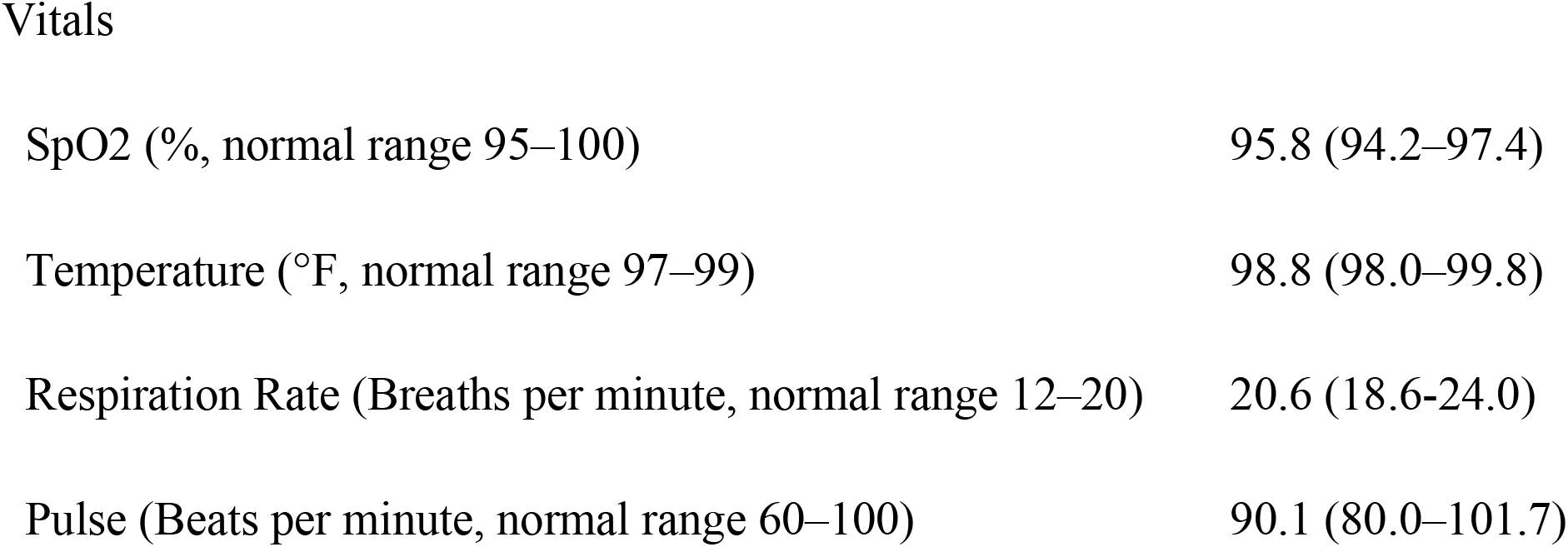
Laboratory measures and vital signs of hospitalized patients on their dates of admission.

| Measures | Median (IQR) |
| --- | --- |
| Laboratory tests |  |
| White Blood Count (K/UI, Low: 4.00, High: 10.00) | 7.3 (5.4–10.0) |
| Neutrophil Absolute Count (K/UI, Low: 1.84, High: 7.80) | 5.2 (3.6–7.7) |
| Lymphocyte Number (K/UI, Low: 0.72, High: 5.20) | 1.1 (0.8–1.5) |
| Monocyte # (K/UI, Low: 0.12, High: 1.00) | 0.5 (0.3–0.7) |
| Neutrophils Percent (% , Low: 46.0, High: 78.0) | 74.6 (65.9–81.7) |
| Lymphocyte Percent (% , Low: 18.0, High: 52.0) | 16.3 (10.6–23.2) |
| Monocyte % (% , Low: 3.0, High: 10.0) | 7.0 (4.9–9.2) |
| Albumin (G/Dl, Low: 3.5, High: 5.0) | 3.3 (3.0–3.7) |
| Sgot (U/L, Low: 3, High: 44) | 38.0 (25.0–59.0) |
| Sgpt (U/L, Low: 0, High: 40) | 30.0 (18.0–49.0) |
| Red Blood Count (M/UI, Low: 4.00, High: 5.20) | 4.4 (3.9–4.9) |
| Urea Nitrogen (Mg/Dl, Low: 8, High: 21) | 14.0 (10.0–24.0) |
| Creatinine (Mg/Dl, Low: 0.65, High: 1.00) | 1.0 (0.8–1.4) |
| Hemoglobin (G/Dl, Low: 12.0, High: 16.0) | 12.9 (11.1–14.2) |
| Ferritin (Ng/Ml, Low: 12, High: 260) | 766.2 (325.9–1643.0) |
| D-Dimer (Mg/L Feu, Low: 0.00, High: 0.60) | 0.9 (0.5–2.5) |
| C-Reactive Protein (Mg/L, Low: 0.0, High: 8.0) | 112.1 (54.5–192.8) |
| Lactate Dehydrogenase (U/L, Low: 110, High: 240) | 404.0 (304.0–544.0) |
| Glucose, Blood (Mg/Dl, Low: 60, High: 99) | 122.0 (103.0–175.0) |
| Platelet Count (K/UI, Low: 150, High: 399) | 218.0 (170.0–281.0) |
| Creatine Phosphokinase (U/L, Low: 10, High: 205) | 128.0 (66.0–332.5) |
| Vitals |  |
| SpO2 (% , normal range 95–100) | 95.8 (94.2–97.4) |
| Temperature (°F, normal range 97–99) | 98.8 (98.0–99.8) |
| Respiration Rate (Breaths per minute, normal range 12–20) | 20.6 (18.6-24.0) |
| Pulse (Beats per minute, normal range 60–100) | 90.1 (80.0–101.7) |

**Table 3.**
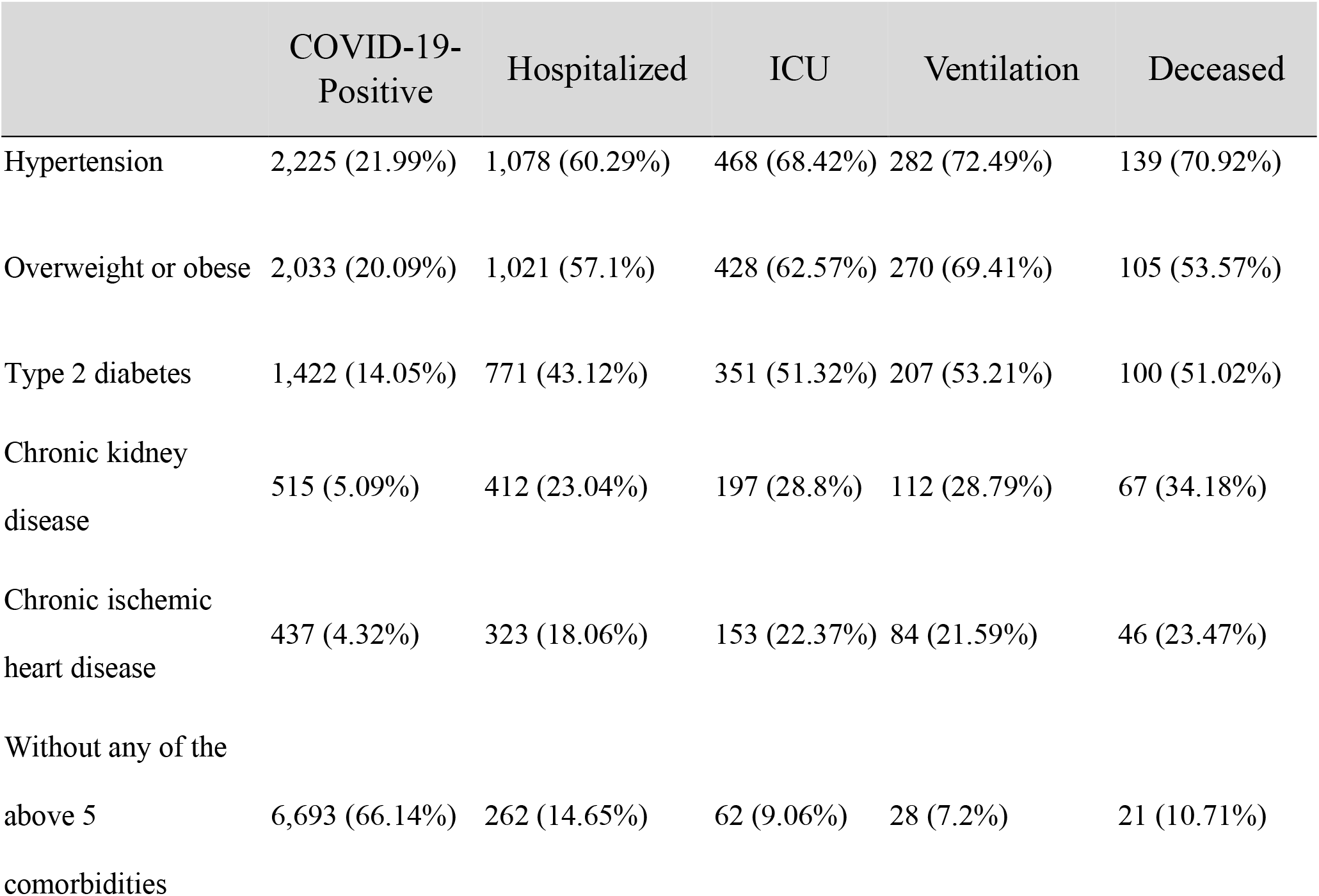
Comorbidities among the COVID-19 patients.

### Variables

The variables investigated in our study include the patients’ demographics, laboratory findings, vitals, and comorbidities as collected in the hospital. Demographic variables include patients’ age, sex, first race, and smoking status. Laboratory findings include white blood count, absolute neutrophil count, absolute lymphocyte count, absolute monocyte count, neutrophils percentage, lymphocyte percentage, monocyte percentage, albumin, aspartate transaminase, alanine transaminase, d-dimer, red blood count, blood urea nitrogen, creatinine, hemoglobin, ferritin, d-dimer, C-reactive protein, lactate dehydrogenase, blood glucose, platelet count, and creatine phosphokinase. Vitals include oxygen saturation (SpO2), body temperature, respiration rate, and pulse. Comorbidities include hypertension, overweight condition or obesity, type 2 diabetes, chronic kidney disease, and chronic ischemic heart disease.

### Definition of Stages

We defined the stages of disease progression based on clinical events; data on such clinical events can be used by hospitals to allocate medical resources. In our study, we defined (1) *hospitalization*, (2) *admission to ICU*, (3) *mechanical ventilation*, and (4) *death* as the advancing stages of COVID-19 disease progression. Figure 2 presents the transition of COVID-19-infected individuals across those stages. As shown in the figure, 10,123 individuals tested positive for COVID-19. Among them, 1,788 were treated in the hospital. The patients who suffered more severe disease progression were treated in the ICU and received ventilatory support, depending on the severity of their disease. 684 (38.3%) of the patients admitted to the hospital were treated in the ICU, and of those in the ICU, 380 (55.6%) received ventilation support. 155 ventilated patients died from COVID-19.

**Figure 2.**
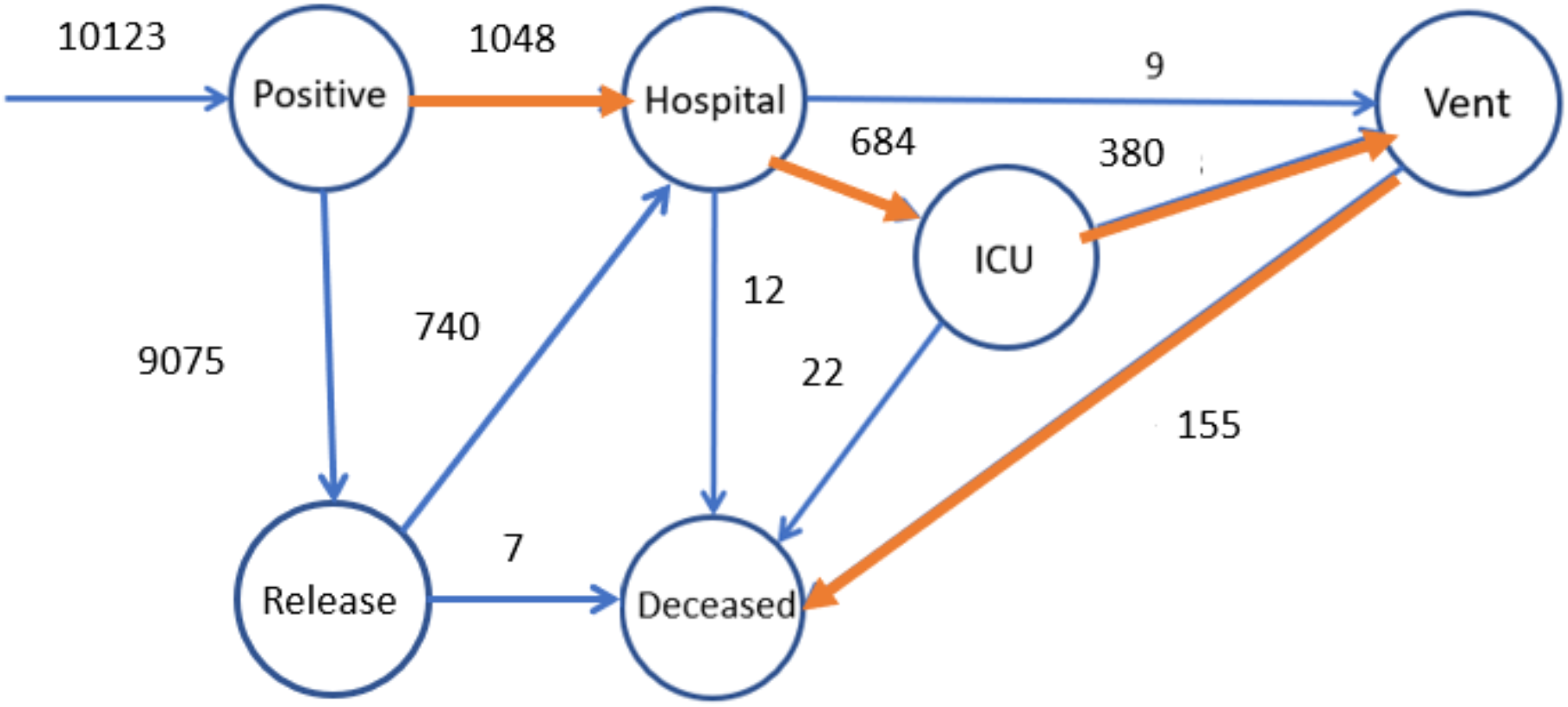
Transitions of patients between stages. 10,123 individuals tested positive for COVID-19. Among them, 1,788 were treated in the hospital. The patients who suffered more severe disease progression were treated in the ICU and some were ventilated, depending on the severity of their disease. 684 (38.3%) of the patients admitted to the hospital were treated in the ICU, and of those in the ICU, 380 (55.6%) received ventilation support. 155 ventilated patients died from COVID-19.

### Analysis of risk factors

LASSO regression [23] was used to select the most discriminative variables for predicting the demand for mechanical ventilation. The “glmnet” packet in R was used to compute the results [24]. A total of 18 variables were entered in an L1-norm LASSO regression. They include age, neutrophil-to-lymphocyte ratio; neutrophil, lymphocyte, and monocyte percentages; white blood, red blood, and platelet counts; and the levels of lactate dehydrogenase, C-reactive protein, blood glucose, blood urea nitrogen, creatinine, albumin, aspartate transaminase, alanine transaminase, hemoglobin, and SpO2. As the regularization term (typically denoted as λ) grows large, only the most important features are left with nonzero coefficients.

In addition to performing a LASSO regression, we also constructed a decision tree [25] for the investigated variables, which. provides another perspective that can be used to identify key risk factors. Using the Gini Index as the impurity metric [26], the decision tree iteratively splits the current data into two branches. By definition, the variable used for splitting the root node is the most discriminative factor. Furthermore, the key risk factors tend to gather at the high-layer nodes near the root. The key risk factors selected by the LASSO regression and decision tree algorithms were further compared and entered in the logistic regression model for predicting ventilator demands.

### Model training and evaluation

We chose to use logistic regression to build the prediction model because an easily interpretable predictive model can help facilitate rapid clinical translation [27]. The key risk factors we identified were used as predictors, and the requirement for ventilation (given as a binary Yes/No variable) was treated as the output label. The task was to predict the demand for mechanical ventilation given data collected at admission. Our positive class (ventilated patients) included 96 patients, and the negative class included 205 patients. The logistic regression classifier was trained and validated using a standard 10-fold cross-validation approach [28]. The accuracy of prediction was evaluated using the area under the receiver-operator characteristic curve (AUC). The mean AUC and confidence intervals were calculated over all cross-validation splits.

## Results

### 1. Overall demographic, symptomatic, laboratory, and vital characteristics

The demographic information, symptoms, and smoking status of infected patients are presented in Table 1. With a median age of 40 and the 75th percentile at 54, the patients were younger than those reported from China [1] and New York [6]. There were more infected females (53.55%) than males (46.45%). In terms of race, African Americans (32.92%) and Whites (27.27%) constituted the largest fractions of the infected patients. Cough (70.14%), fever (46.86%), and shortness of breath (41.32%) were the most common symptoms. In terms of ethnicity, marginally more Hispanics or Latinos (50.88%) were infected than Non-Hispanics or Latinos (49.12%). People who had never smoked (76.42%) and former smokers (16.18%) accounted for most of the infected patients; Current smokers accounted for only 7.4%. The ages of the infected patients are presented in Figure 3(a), which represents data from 03/17/2020 and after. The interquartile range (IQR) of age is represented by the shaded region. The median age decreased from 46 (in week 0) to 37 (in week 20), indicating the spread of disease to a younger population. The number of clinical events, including hospital admissions, ICU admissions, and mortalities, are presented in Figure 3(b). The peak of hospital admissions was reached at week 5; however, the peak of ICU admissions was reached at week 1. The peak of mortality was reached at week 7. Week 7 (which ended on 05/05/2020) was a turning point in the number of hospital admissions, indicating a reduction in transmission of COVID-19 in the Chicago area. However, a second wave of infections is suggested by the increase in hospital admissions from week 14 to week 19.

**Figure 3.**
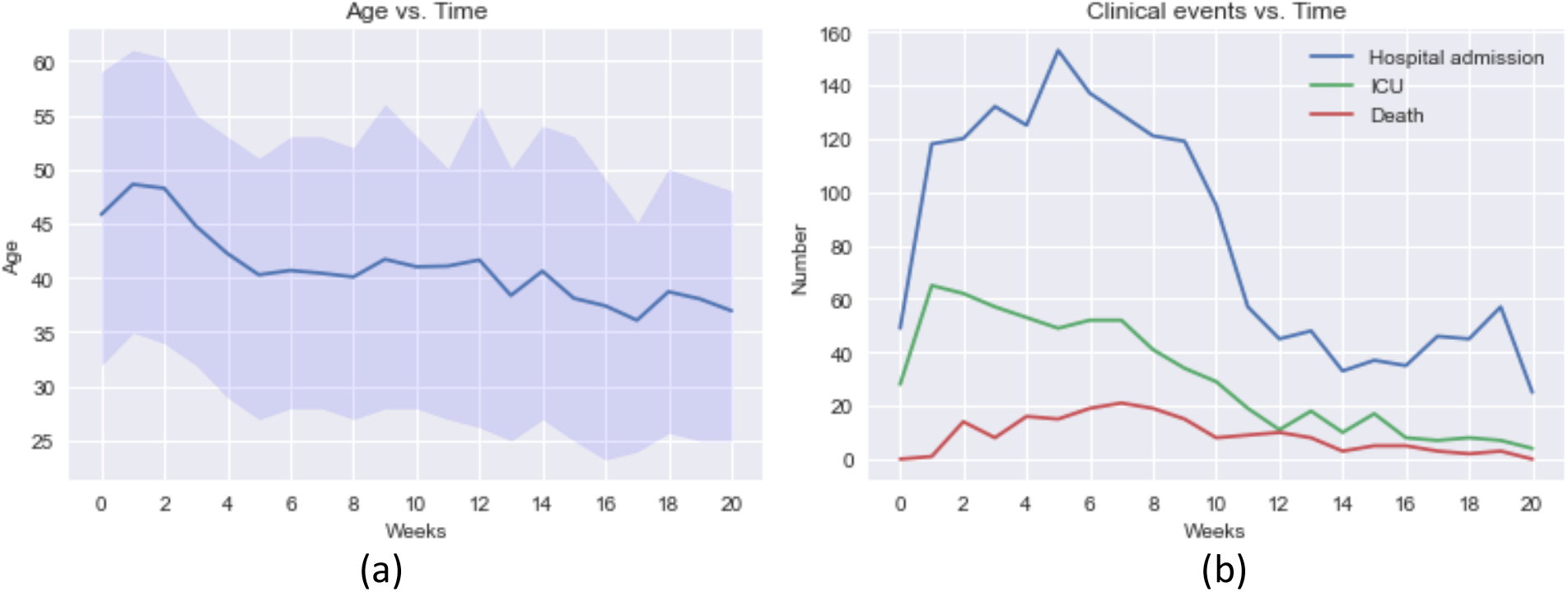
Time-series plot on weekly granularity (data from 03/17/2020 onwards): (a) Age of infected population, (b) Number of hospital admissions, ICU admissions, and deaths.

Table 2 presents the laboratory measures and vitals of hospitalized patients on the day of admission. Among all the investigated laboratory measures, the lymphocyte percentage (median 16.3, IQR 10.6–23.2) and albumin (median 3.3, IQR 3.0–3.7) were lower than the normal range, while ferritin (median 766.2, IQR 325.9–1643.0), d-dimer (median 0.9, IQR 0.5–2.5), C-reactive protein (median 112.1, IQR 54.5–192.8), lactate dehydrogenase (median 404.0, IQR 304.0–544.0), and blood glucose (median 122.0, IQR 103.0–175.0) were higher than the normal range. Among vital signs, the respiratory rate (median 20.6, IQR 18.6–24.0)) was slightly higher than the normal range.

Figure 4 presents the temporal changes in laboratory features, covering 15 days after hospital admission. To reduce the appearance of the same patients in multiple groups, we classified the patients into four groups: hospitalized but not in the ICU; in the ICU but not ventilated; ventilated but not deceased; and deceased. The albumin level of all patient groups decreased from day 0 to day 7. During that time, the albumin levels of the ventilated patients who ultimately survived behaved much like those patients who subsequently died. However, the albumin levels of the eventual survivors then began to increase, while the albumin levels of those who subsequently died continued to decrease until day 11. The blood urea nitrogen level of the patients who eventually died kept increasing until death and was generally higher than the level found in other groups of patients. The lymphocyte percentages of ICU-but-never-ventilated patients tended to move from the abnormal range (<18%) to the normal range, while the lymphocyte percentages of patients who eventually died tended to worsen. That correlation means that the lymphocyte percentage can be used as an indicator of a patient’s condition. A rising lymphocyte percentage indicates a recovery trend in a COVID-19 patient, while a decreasing lymphocyte percentage indicates a worsening condition. As for the red blood count, the initial values for patients who later died were in roughly the same range as those of survivor groups those of survivor groups, but later the value for patients who died showed a sharper decrease compared with the survivors. Thus, the initial value for the red blood count may not be a risk factor for mortality prediction, but its downward trend can be used to forecast deterioration.

**Figure 4.**
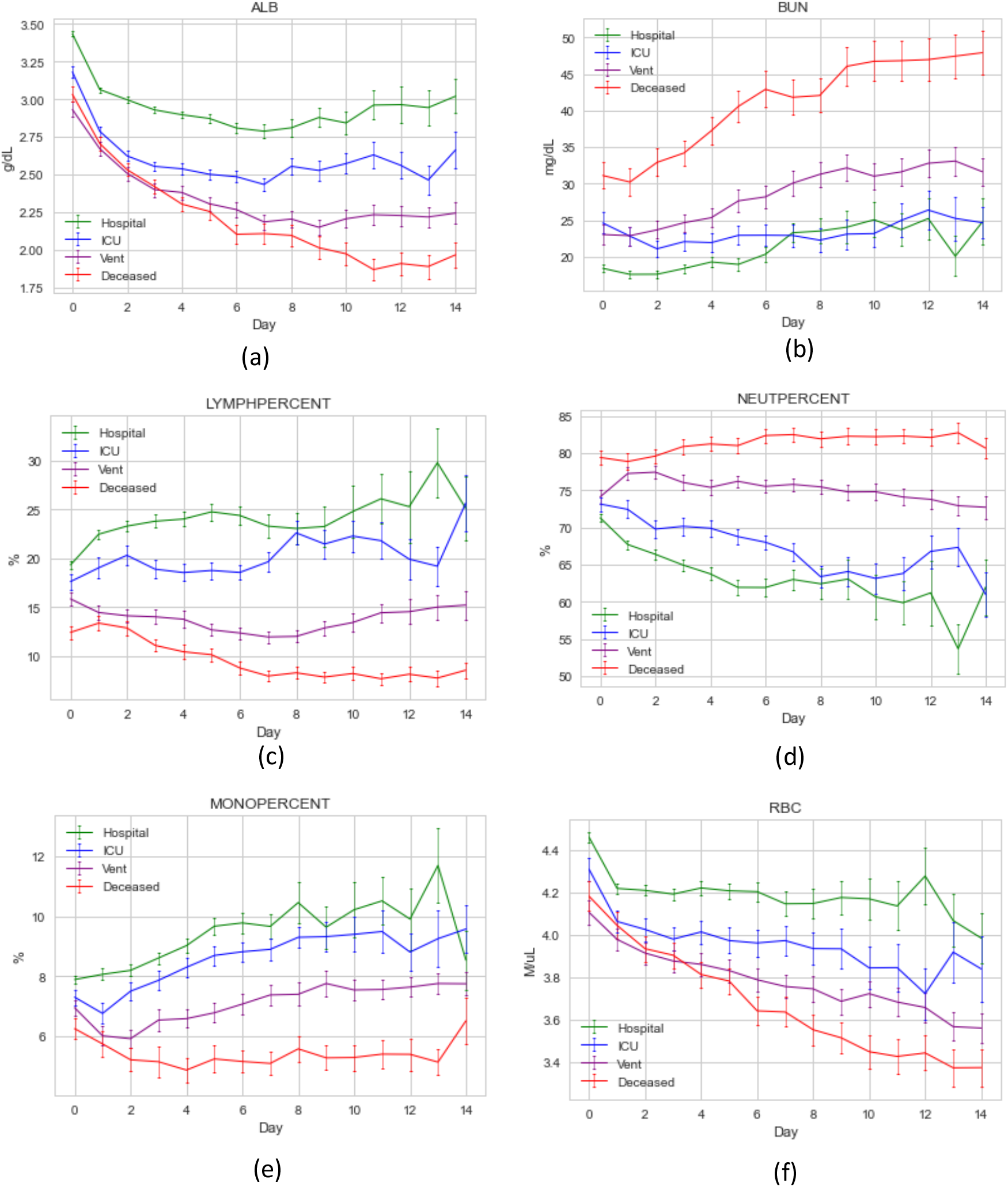
Temporal changes of laboratory features from the day of hospital admission to 15 days in hospital: (a) albumin level, (b) blood urea nitrogen level, (c) lymphocyte percentage, (d) neutrophil percentage, (e) monocyte percentage, (f) red blood count.

### 2. The impact of comorbidities on disease progression

We investigated five comorbidities for COVID-19 patients. Ranked by their rates of appearance among the hospitalized patients, they include hypertension (60.29%), overweight or obese condition (57.1%), type 2 diabetes (43.12%), chronic kidney disease (23.04%), and chronic ischemic heart disease (18.06%). As COVID-19 progressed from the hospitalization stage to the ICU stage to eventual death, the percentage of patients who had chronic kidney disease or chronic ischemic heart disease constantly increased, indicating that these two comorbidities are significant risk factors for severe disease progression. In addition, the rates of hypertension and type 2 diabetes were also higher among deceased patients than among the total hospitalized population. The patients who had none of these five comorbidities and presented as relatively healthy accounted for very few (14.65%) of the hospitalized patients and even fewer of the deceased patients (10.71%), meaning that an originally healthy individual is far less likely to suffer serious effects due to COVID-19. These findings indicate that the studied comorbidities are risk factors for mortality.

### 3. Risk factors of severe disease progression and predicting ventilation demand

Figure 5(a) shows the trace of coefficients as the λ grows large. The coefficients of investigated features turn to 0 sequentially. Among all the features, albumin is the last one that turns to 0, meaning that albumin is the most discriminative feature selected by LASSO, followed by SpO2 and white blood count. The significance of albumin is also indicated by the magnitudes of coefficients; the coefficient of albumin is consistently larger than that of any other feature. Albumin can be considered a general measure of an individual’s overall health. Figure 5(b) shows the trace of binomial deviance, a type of misclassification error [29]. As the regularization parameter λ becomes large, the binomial deviance first decreases and then increases. The axis above the figure shows the number of nonzero coefficients at a particular λ value. The minimal deviance is achieved with 12 features. They include age, platelet count, white blood count, neutrophil-to-lymphocyte ratio, lymphocyte percentage, and the levels of lactate dehydrogenase, C-reactive protein, blood glucose, blood urea nitrogen, albumin, hemoglobin, and SpO2.

**Figure 5.**
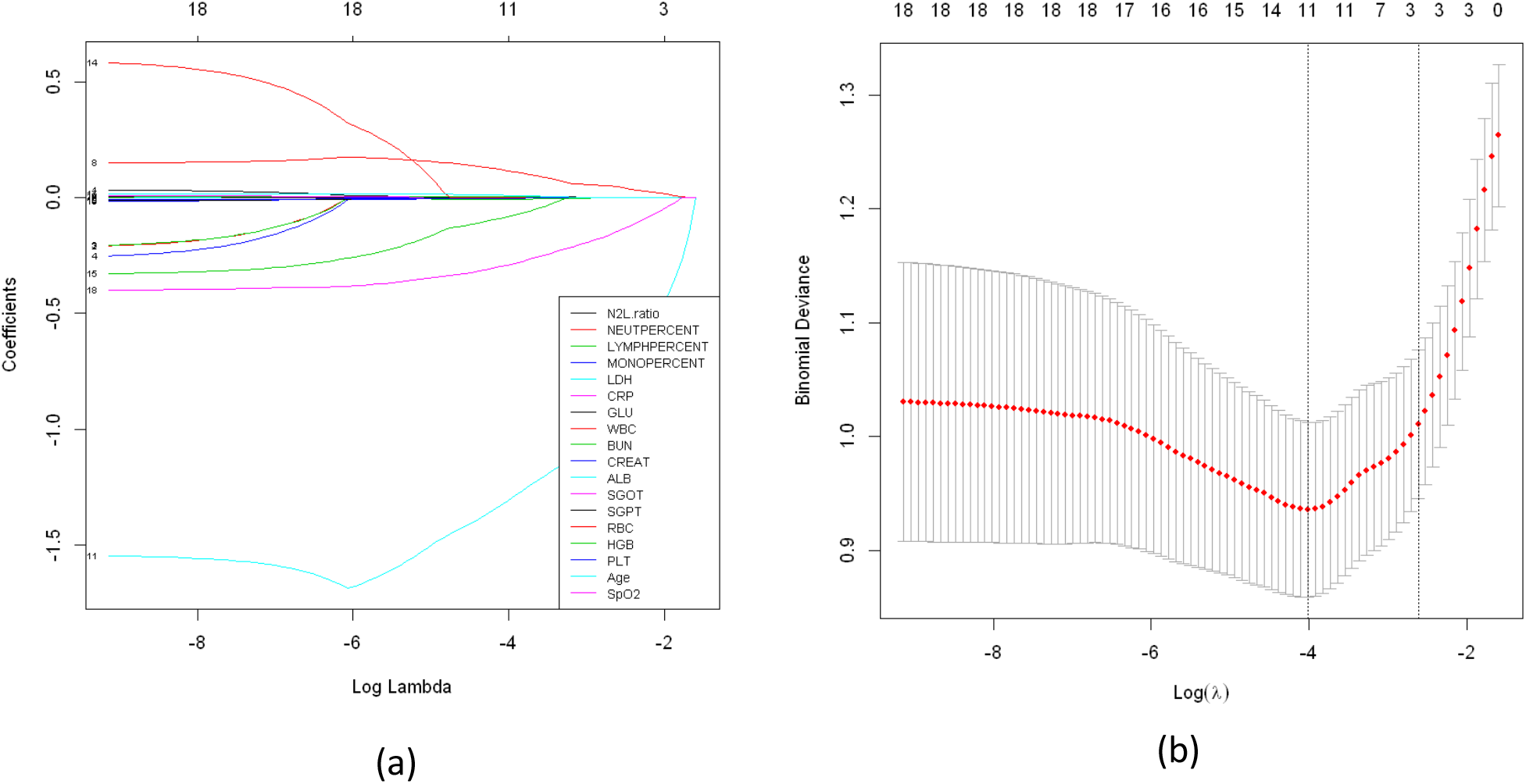
LASSO regression for feature selection. (a) Trace of coefficients of the 18 baseline features. (b) Binomial deviance using 10-fold cross-validation, indicating the variation in misclassification error with different levels of regularization.

Using the 12 features selected by LASSO regression, we achieved an AUC of 0.819 (95%CI: 0.751–0.888) using a logistic regression approach. To identify the most discriminative features, we used the decision tree algorithm with the Gini Index. Figure 6 shows the result of the decision tree with max layer = 3. The first split is made on albumin = 2.65 g/dl. For the patients with albumin lower than 2.65 g/dl, the second split is made on the white blood count = 6.835 k/ul, indicating that the patients with low albumin and high white blood count are more likely to require mechanical ventilation. Whereas, for the patients with albumin higher than 2.65 g/dl, the second split is made on SpO2 = 92.739%, indicating that the patients with high albumin and high SpO2 values are less likely to need ventilatory support. The top three features selected by the decision tree algorithm exactly match those selected by LASSO regression, highlighting the significance of these features. In clinical practice, this compact set of features may be used to efficiently triage COVID-19 patients. We trained another logistic regression model with these three features alone and achieved an AUC of 0.823 (95%CI: 0.765–0.880), which is marginally higher than the AUC achieved using all 12 features.

**Figure 6.**
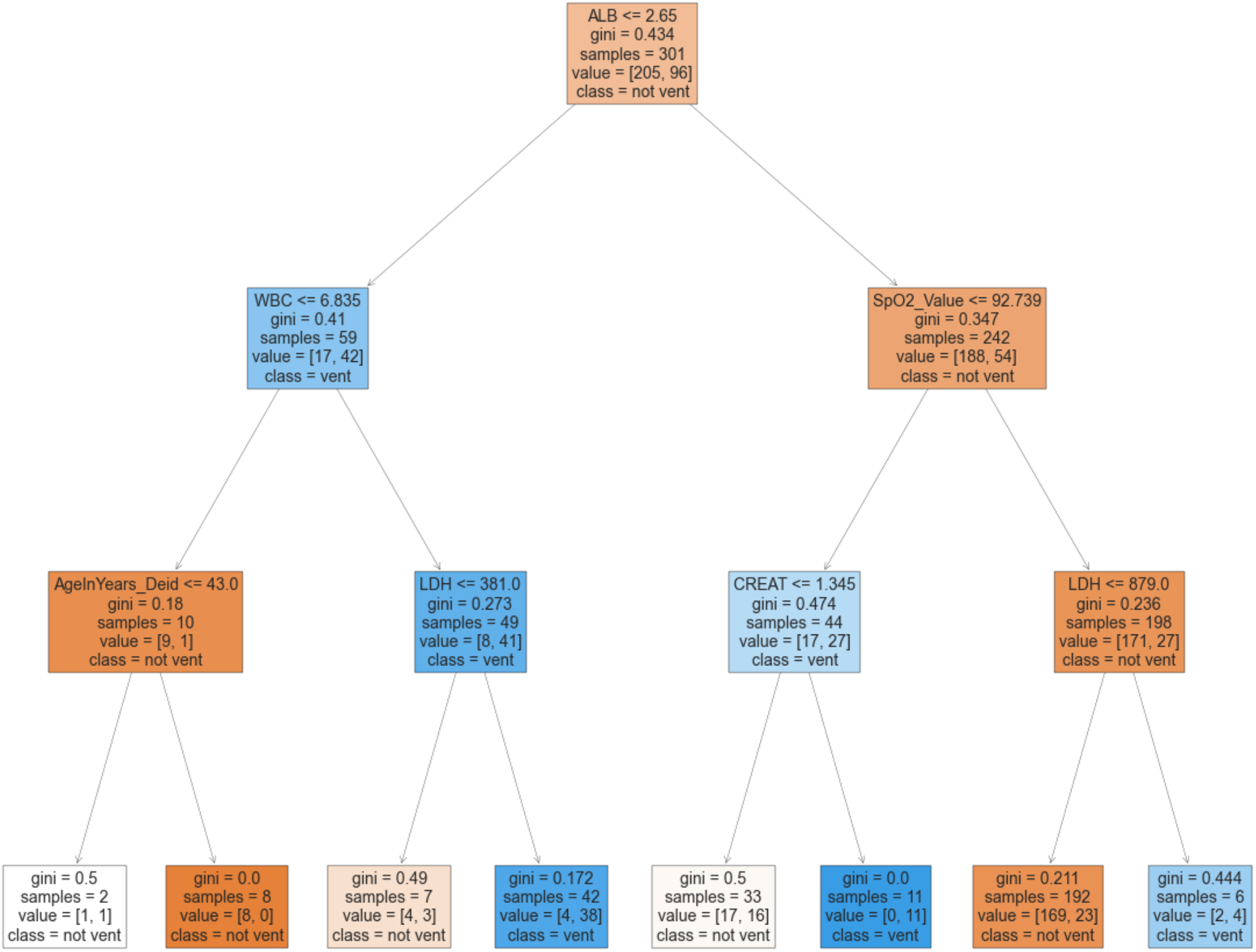
Decision tree for distinguishing between patients who did and did not require ventilation. A maximum tree depth of 3 and the Gini impurity index were utilized to derive the decision tree. The first split is made on albumin = 2.65 g/dl. For the patients with albumin lower than 2.65 g/dl, the second split is made on the white blood count = 6.835 k/ul. Whereas, for the patients with albumin higher than 2.65 g/dl, the second split is made on SpO2 = 92.739%.

We selected a cutpoint of the prediction model by maximizing the sum of sensitivity and specificity, which is also formally equivalent to maximizing Youden’s Index [30]. Such a criterion is desirable in clinical setting for it maximizes the overall correct classification rate and thus reduce the overall misdiagnosis rate. We performed a grid search by varying the cutpoint from 0 to 1 with an interval of 0.05. The maximal sum of sensitivity and specificity was achieved at the cutpoint of 0.35, where sensitivity = 0.70 and specificity = 0.82.

## Discussion

This paper presents a novel approach for triaging COVID-19 patients in a hospital setting to predict the need for mechanical ventilation. We created the approach using a multi-stage perspective in a retrospective manner. Our data are unique in that they came from a large metropolitan hospital with an extremely diverse patient population. We characterized the stratification of COVID-19 patients in terms of clinical events and presented laboratory measures in both static and dynamic views, thereby making it easier to understand and track disease progression. We used LASSO regression and decision tree models to identify several risk factors for deterioration of patient health. Using those identified risk factors, we developed a logistic regression-based prediction model that can identify patients at risk of clinical deterioration. With a cutpoint of 0.35, the model performed well with a positive predictive value of 0.644. At Rush, the use of this model would have correctly identified 67 (out of 96) patients who would require ventilation support, with 37 false positives (out of the 205 patients who did not require ventilation support).

This research and the resulting model establish the feasibility of an early triaging tool that can predict the clinical course of COVID-19 at a subject-specific level. This not only has the potential to improve patient outcomes, but also can provide hospital administrations with insights that inform resource management.

Several other previously published modeling approaches have attempted to evaluate predictors of clinical deterioration, mechanical ventilation, and death. Factors associated with disease progression in non-peer-reviewed work include lower platelet and lymphocyte counts; increased markers of DIC, such as fibrinogen, d-dimer, and aPTT; increased LDH, AST, and CK, and abnormal CT scans [31]; clinical comorbidities, CRP, respiratory rate, and LDH [32]; and higher SOFA score, age, and d-dimer levels [33]. One non-peer-reviewed study found similar factors and developed a nomogram for prediction [34]. In a peer-reviewed study, clinical comorbidities were the most predictive for severe disease progression [35]. Our work has identified three critical indicators of the deterioration of COVID-19 patients, namely albumin, SpO2, and white blood count, which are readily measurable in a clinical setting.

Future work with this model will implement an early warning system for clinical disease progression. The value of using an approach that recognizes respiratory compromise is that it can identify patients for whom early intervention can improve outcomes. With the insights gleaned from emerging clinical data, the use of optimized prone positions, medical therapy with antivirals, and anti-inflammatory medication may abbreviate the inflammatory response, improve oxygenation, reduce the risk of intubation, and reduce mortality in patients with COVID-19. An approach like the one introduced in this paper can also identify patients for whom early discharge is safe. A triage tool for sorting high- vs. low-risk individuals with COVID-19 would be highly useful in resource-constrained situations in which bed capacity must be tightly managed.

Our study is subject to several limitations. It is a single-center study, and practice patterns in that center may not generalize to other centers. In addition, we did not include radiography in the predictive model. Future studies will continue refining the model and validate its performance prospectively.

## Data Availability

The data is not publicly available.

## Acknowledgements

We thank Kathleen Atchley and Jenny Applequist for assisting with preparing the manuscript. This project has been funded by the Jump ARCHES endowment through the Health Care Engineering Systems Center at the University of Illinois.

## Notes

### Competing Interest Statement

The authors have declared no competing interest.

### Author Declarations

Rush University Medical Center

